# Effect of maximum voluntary isometric contraction of the triceps surae muscle on a subsequent drop jump

**DOI:** 10.1101/2024.09.04.24312972

**Authors:** Taiki Ino, Keita Ogawara, Shuichi Uchiyama, Shinya Endo, Seiji Miyazaki, Hiroshi Yamada

## Abstract

**Purpose:** Pre-muscle contraction improves sports performance because post-activation potentiation (PAP), induced by a previous intense voluntary contraction (conditioning contraction), amplifies the subsequent target muscle contraction. This study aims to examine the influence of conditioning contraction on the performance of ballistic motion, particularly on the stretch-shortening cycle (SSC).

**Methods:** Fourteen male university students specializing in athletic jumping events performed a drop jump from a height of 0.3 m. Maximal voluntary isometric plantarflexion for 6 seconds was considered a conditioning contraction. We set two conditions: in the PAP condition, participants performed a conditioning contraction 10 s before the drop jump, and in the control condition, they simply performed the drop jump. After 10 minutes of rest from the reference drop jump, both conditions were performed. A 3D motion analysis system, force plates, and surface electromyography were used to record the jump data.

**Results:** In the PAP conditions, the jump height and velocity of the center of gravity increased by 4 cm and 14 cm/s, respectively. Lower limb torque increment was observed only in the ankle joint between the PAP and control conditions (0.31 Nm/kg). However, there was no change in the magnitude of muscle activity based on the electromyogram.

**Conclusion:** Our study showed that PAP could potentiate the contraction mechanism under excitation-contraction coupling, irrespective of the effect on the central nervous system. Therefore, PAP enhances jump performance by improving SSC.

## INTRODUCTION

In competitive sports, explosive power is required in many situations, such as the takeoff motion in athletics and volleyball spike jump competitions. This explosive power is generated by the stretch-shortening cycle (SSC), a shortening motion immediately after impulsive stretching. The main SSC mechanism for jumping motions is that muscle groups and tendons in lower limb joints absorb and reuse kinetic energy as elastic energy. SSC shortens the ground contact time of high jumping motions, such as rebound jumps;^1,2^ particularly, the SSC occurs more commonly in the ankle joints than in the knee and hip joints. Regarding structure, the ankle joints produce more torque induced by eccentric contraction compared to other joints.^3,4^ Additionally, the ankle joints are significant in different jumping motions.^5^ This ankle movement during a jump acts like a viscoelastic body because of the stiffness of muscle fibers (number of joined cross-bridges and nature of myofibrils), tendons, membrane, and pennate muscles.^6,7^ Thus, increasing the magnitude of muscle contraction in the triceps surae muscle improves SSC capability, which is vital for jumping motions performed under high momentary loads.

Post-activation potentiation (PAP) is a phenomenon in which voluntary exercises increase the muscle strength exerted during subsequent exercises.^8^ Its mechanism of action is muscle contraction rather than excitation of the nervous system or the peripheral nervous system. Muscle contraction is the sliding of myosin and actin filaments, controlled by the concentration of calcium ions. Calmodulin, activated by binding with calcium ions, activates myosin light chain kinase, and the activated myosin light chain kinase phosphorylates the myosin light chain.^9^ Therefore, actomyosin becomes more susceptible to binding to actin, actomyosin sensitivity to calcium ions increases, and muscle contraction increases.^10^ This mechanism enables PAP to improve muscle strength. However, the rest time and magnitude of conditioning contraction (CC), a voluntary exercise for inducing PAP, often performed at a higher intensity than subsequent exercises, critically influence the enhancing effect. A study using resistance training events and maximal voluntary isometric contractions for CC reported that 4 minutes of rest did not improve performance.^11^ An increase in voluntary muscle strength was observed immediately after a maximal voluntary isometric contraction for 6 seconds without a rest period.^12^ Furthermore, a study comparing the effect of maximum voluntary isometric contractions of 3 sets for 3 seconds versus 3 sets for 5 seconds reported that 3 sets for 3 seconds improved jump performances, such as drop jump but 3 sets for 5 seconds decreased them due to fatigue.^13^ PAP is ineffective for improving performance because high-intensity CC causes fatigue while inducing greater enhancement; thus, rest is required before the subsequent exercise. Because components of SSC such as stretch-reflex and muscle stiffness deteriorate with muscle fatigue,^14^ minimizing fatigue and utilizing the enhancement is needed for observing the effect of PAP on SSC. Some studies manipulate the duration of CC to induce PAP with less fatigue.^12,13,15^ However, no agreement has been reached because few studies applied immediate PAP without a rest period to instantaneous and explosive force exertion.

Because of these inconsistencies, the rest period for CC, the choice of CC training, and the individual differences observed were most likely to blame. When using high-intensity resistance training like CC, it is difficult to establish rest periods to maintain a balance between the effects of fatigue and the effect of PAP on subsequent exercises.^16-18^ Furthermore, many technical factors are involved in multi-joint movements, such as resistance training. Individual differences were observed in the magnitude of stimulation of target muscles. Some conditions are required for CC, which is convenient and effective for exerting PAP and appropriate for observing the effect; it causes less fatigue, does not require a set rest period, and is free of technical factors. Therefore, the CC was found to be an isometric maximum voluntary contraction that requires no technical factors and induces PAP with no rest period. CC could potentiate a drop jump in which the SSC movement occurs in the ankle joints.

This study aimed to examine the effects of isometric CC on the drop jump, investigate the effect of PAP on the SSC, present useful findings for further research, and suggest preparatory movements that improve performance immediately.

## METHODS

### Participants

The participants were athletes used to jumping with high loads for the following reasons: if the stretching load is too large for people who are not used to plyometrics, the jump movement becomes a protective movement for the muscle-tendon complex, and the SSC cannot function properly.^19^ Furthermore, when the heel touches the ground, the excitability of the plantarflexion muscles is suppressed, the elastic energy storage-reuse mechanism does not function effectively, and the transition from stretching to shortening takes time.^20,21^ Thus, 14 males (age: 21.1 ± 1.4 years, height: 173.6 ± 4.7 cm, weight: 64.7 ± 4.0 kg, long jump record: 7.32 ± 0.32 m) from the university athletics club jumping event who regularly perform jump training such as drop jump volunteered to participate. The target leg was determined to be the takeoff leg for a long jump. This study was approved by the Ethics Committee of Tokai University for “Research on Human Subjects” (approval number: 16096) and was conducted after all the participants provided informed consent.

### Trial

The trial consisted of a drop jump from a 0.3-meter-high platform and a jump up. This attempt was suitable for evaluating the ballistic movement.^22^ The trial was performed three times in a row under every condition in the case of failure. The trial did not induce a potentiation effect for subsequent trials.^16,23^ The participants were instructed to put their takeoff leg forward and then fall without jumping off the platform, to control their arms’ motion by placing their hands on their waist during the trial, and to jump barefoot. The participants were instructed to “shorten the contact time as much as possible and jump straight up at full power”.

### Conditions

Two conditions were set: the PAP condition with intervention and the control condition. Participants performed the drop jump twice in both conditions, pre-intervention (pre) and post-intervention (post), with a 10-min rest in between. CC was performed 10 s before post only in the PAP condition. The maximum voluntary isometric plantar flexion was performed for 6 s as CC, which can induce PAP immediately.^12,15^ Therefore, the 10 s duration between CC and the trial was set to equalize each attempt. The ankle joint was positioned at 90° with the participant sitting forward.

### Procedure

Figure 1 shows the study procedure. During this trial, the participants initially practiced extensively. After a 10-minute rest from practice, the PAP or control condition was randomly performed. The rest time between pre and post was 10 minutes, which eliminated the influence of PAP in this CC.^12,15^ Twenty minutes after finishing either condition, another condition was performed. At rest, the participants were placed in a sitting position to suppress physical activity as much as possible.

**Figure 1:**
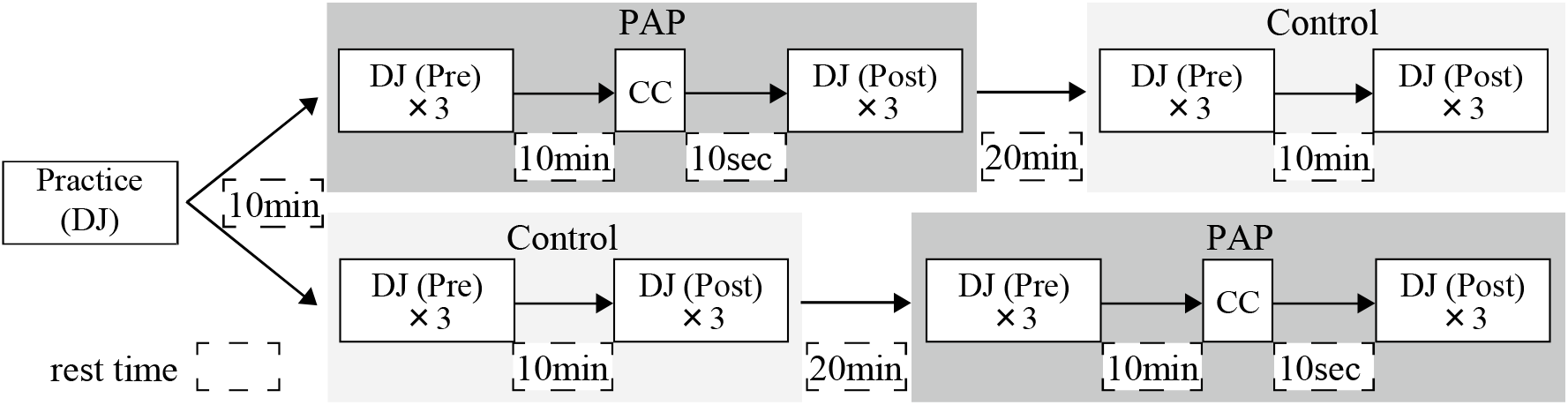
The study procedure. Participants performed both PAP and control conditions with the upper or lower procedure in the figure. PAP: PAP condition, Control: control condition, DJ: drop jump, CC: conditioning contraction (maximal voluntary isometric plantarflexion for 6 seconds), Pre: preintervention trial, Post: post-intervention trial.

### Measurement and analysis methods

All trials were measured and calculated using the 3D coordinates of the 32 markers and a 3D motion analysis system (Mac3D, manufactured by Motion Analysis). The frame rate and shutter speed were 250 s and 1/500 s, respectively. The markers were attached to the participant’s body surface using the Japanese model for estimating inertia properties.^24^ The angles and angular speeds of the hip, knee, and ankle joints were calculated from the 3D coordinates. Each segment’s coordinates and inertial moments were estimated from the model and calculated using the floor reaction force data. A 3D video motion analysis system (Frame-DIAS V; DKH) was used to calculate the items.

Two force plates (FP6090-15, FP4060-15, Bertec) with a sampling frequency of 1,000 Hz was used to record the ground reaction force.

An electromyogram (EMG) was derived and recorded using surface electromyography with bipolar electrodes (DL-141, S & ME), with an inter-electrode distance of 12 mm, from the tibialis anterior (TA), soleus (SOL), medial gastrocnemius (MG), lateral gastrocnemius (LG), rectus femoris (RF), and biceps femoris (BF). The sampling frequency was 1,000 Hz. The root mean square (RMS) of EMG in each phase was calculated, and muscle activity was normalized (%RMS) by dividing the RMS by the muscle activity of the maximum voluntary contraction.

### Phase definition

Ground contact and takeoff were determined by the vertical ground reaction force, and the analysis range was set from ground contact to takeoff. The vertical displacement of the synthetic center of gravity of the body in the analysis range was calculated from the motion analysis data to determine the transition point between descent and ascent. Based on the above data, the two phases were determined to be the descending phase (ground contact–transition point) and the ascending phase (transition point–takeoff).

### Statistical analyses

Statistical analyses were performed using R (version 2.14.1, platform i386-apple-dorwin 9.8.0). The target trial for the analysis was the first drop jump in a trial, unless it failed. A two-way repeated-measures analysis of variance was performed to observe the impact of CC on jump height, the velocity of the center of gravity at takeoff, lower limb joint torque, ground contact time, ground reaction force impulse, and % RMS. For items with an interaction, a paired t-test was used within each trial (pre- and post-intervention and between conditions) and considered significant at *P* < .05. Results were reported as means and standard deviation (SD).

## RESULTS

### Jump height

The jump height was evaluated to determine the drop jump’s performance. Figure 2 shows the pre- and post-jump heights to determine the drop jump in each condition. An interaction was observed between the control and PAP conditions (*P* = .0010). When comparing conditions in each trial, jumping height under the control and PAP conditions was not different pre-test (*P* = .4103). However, the PAP condition showed a significantly higher value post-test (*P* = .0044). Furthermore, when comparing pre- and post-exercise under each condition, there was no change in the control condition (*P* = .7475). However, there was a significant improvement post-exercise under the PAP condition (*P* = .0003). In the control condition, pre was 0.362 ± 0.036 m, and post was 0.360 ± 0.044 m. In the PAP condition, pre was 0.354 ± 0.042 m and post was 0.393 ± 0.042 m, showing an 11% increase of 0.039 m.

**Figure 2:**
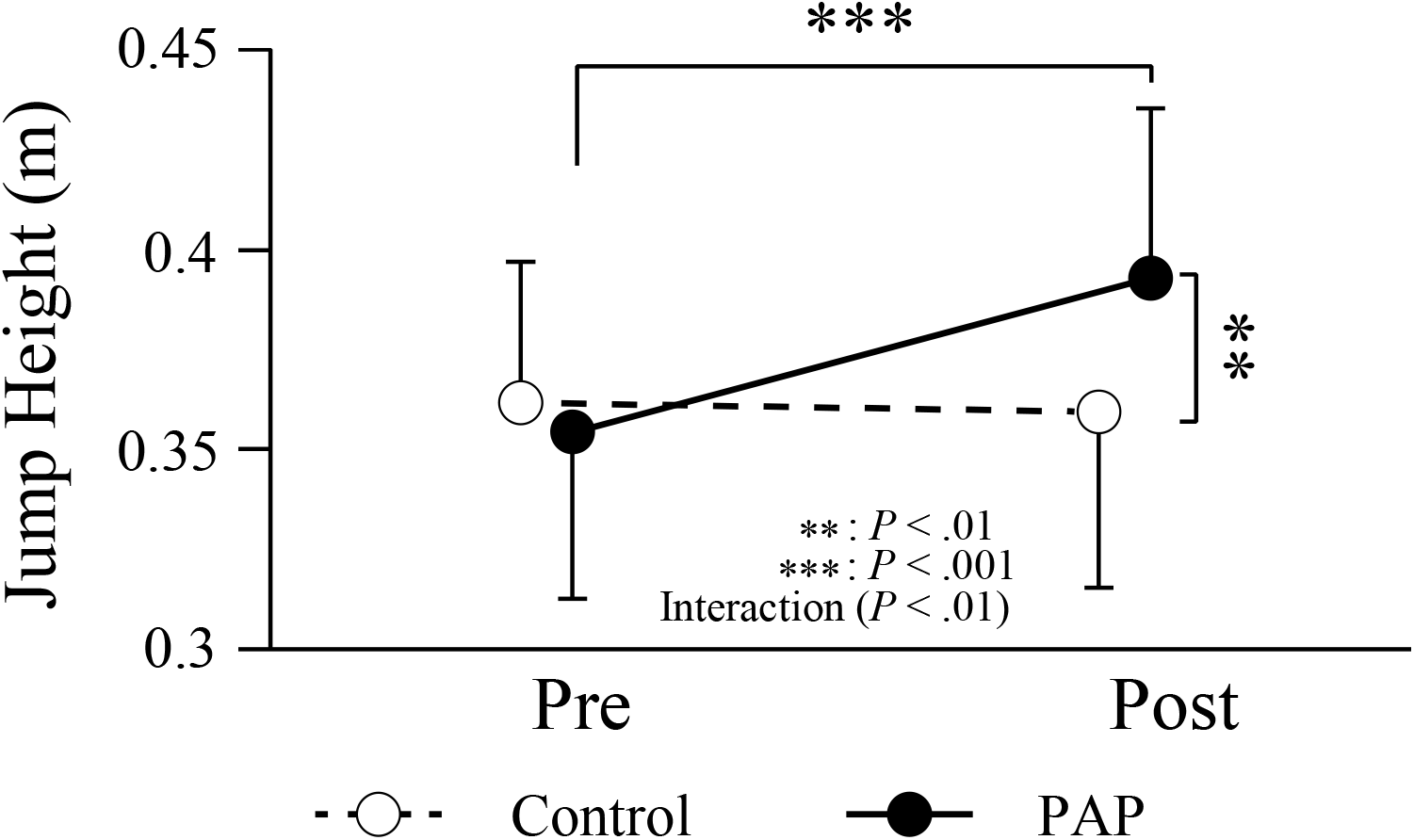
Jump height of drop jump in pre and post in PAP and control condition. Circles and error bars indicate means and SDs respectively. Black circle with black line illustrates the PAP condition. White circle with dashed line shows control condition.

### Velocity of the center of gravity at takeoff

Because jump height is influenced by the initial velocity of the body at takeoff, it was viewed as an important factor of a jump performance. Figure 3 shows the velocities of the center of gravity at takeoff under each condition. An interaction was observed between the control and PAP conditions (*P* = .0019). Comparing the conditions in each trial, post under the PAP condition showed a significantly higher value than that in the control condition (*P* = .0118), while, pre was not significantly different (*P* = .4506). Additionally, when comparing pre- and post-intervention in each condition, there was no change in the control condition (*P* = .7252), but there was a significant improvement post-intervention in the PAP condition (*P* = .0021). In the control condition, pre was 2.52 ± 0.19 m/s and post was 2.50 ± 0.20 m/s. Under the PAP condition, pre was 2.49 ± 0.20 m/s and post was 2.63 ± 0.18 m/s, showing an increase of 0.14 m/s.

**Figure 3:**
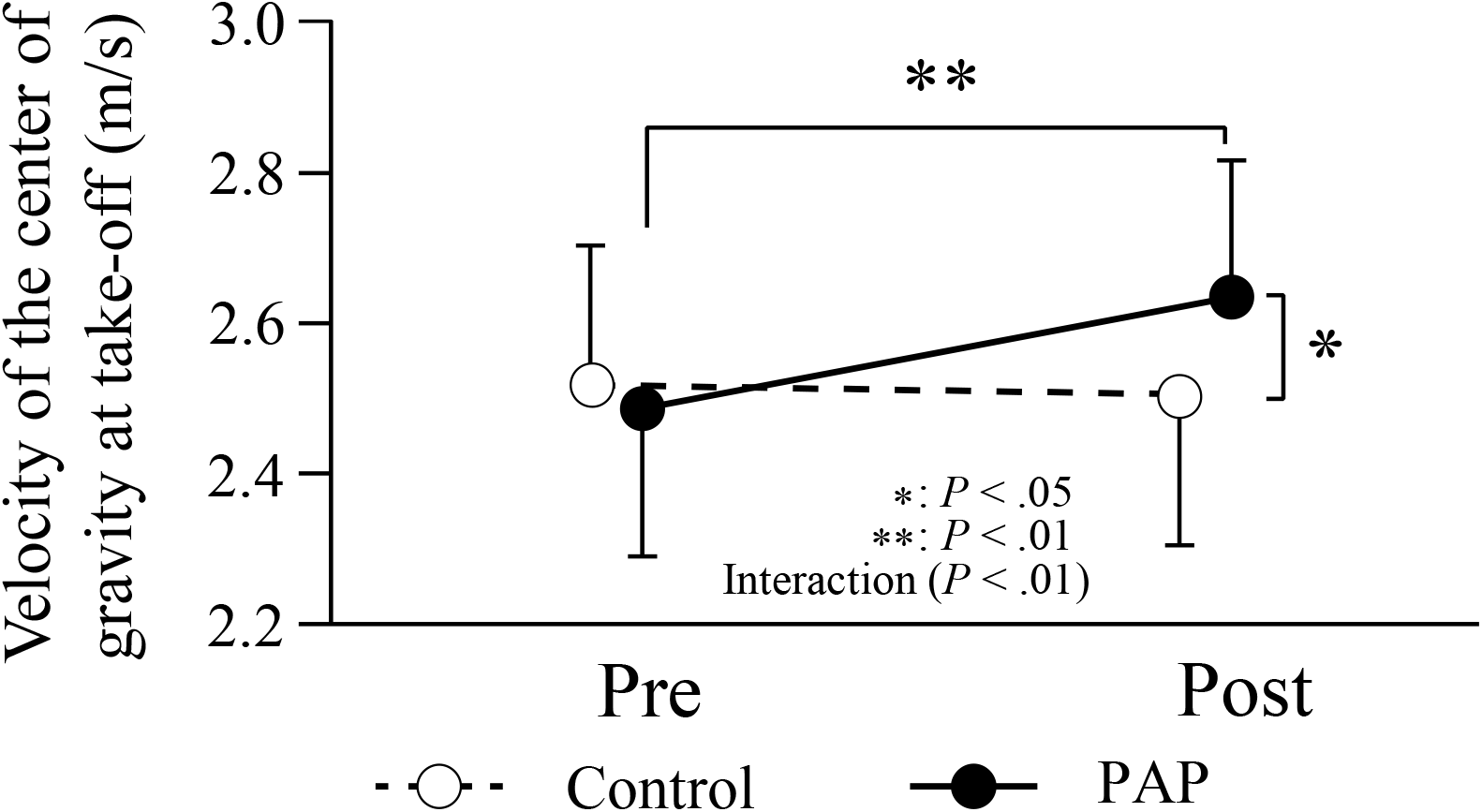
Velocity of the center of gravity at takeoff in pre and post in PAP and control conditions. Circles and error bars indicate means and SDs respectively. Black circle with black line illustrates PAP condition. White circle with dashed line shows control condition.

### Ground contact time

No significant interaction was observed between conditions (*P* = .0751). In the control condition, pre was 0.16 ± 0.03 seconds, and post was 0.17 ± 0.02 seconds. Under the PAP condition, pre was 0.16 ± 0.03 seconds and post was 0.16 ± 0.03 seconds, and there was no difference between them. According to the reports about drop jump, the ground contact time is within approximately 0.2 seconds,^25-27^ all trials were performed with a reasonable and appropriate ground contact time.

### Ground reaction force impulse (vertical)

Figure 4 shows pre- and post-test of impulse responses under each condition. An interaction was observed between the control and PAP conditions (*P* = .0040). Comparing the conditions in each trial showed that the PAP condition was significantly higher only in post (*P* = .0105) but no significant difference in pre (*P* = .3798). Furthermore, there was no change in the control condition when comparing pre- and post-intervention (*P* = .2797). However, there was a significant improvement in the PAP condition (*P* = .0043). Under the control condition, pre was 228 ± 24 N s, and post was 224 ± 21 N s. Under the PAP condition, pre was 225 ± 24 N s and post was 236 ± 23 N s, showing an increase of 11 N s.

**Figure 4:**
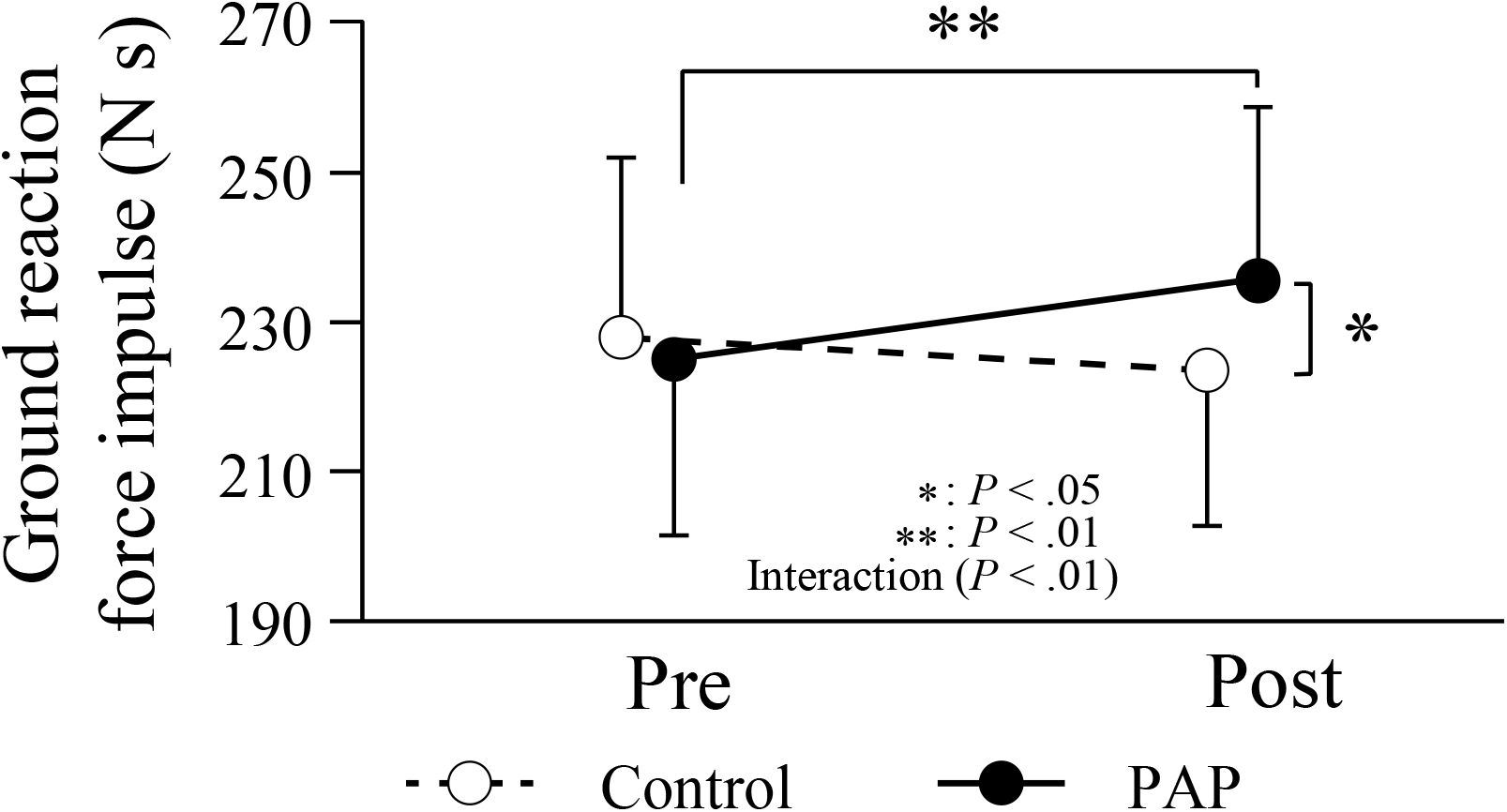
Ground reaction force impulse in pre and post in PAP and control condition. Circles and error bars indicate means and SDs respectively. Black circle with black line illustrates PAP condition. White circle with dashed line shows control condition.

### Joint torque (lower limb)

No significant interaction was observed between the control and PAP conditions in the knee and hip joints torque (*P* = .416 and .0791, respectively). However, there was an interaction in the ankle joint (*P* = .0407). Figure 5 shows the maximum ankle joint torque pre- and post-test under each condition normalized by dividing by the participant’s weight. When comparing trials between conditions, the PAP condition showed a significantly higher value in post (*P* = .04320) but not in pre (*P* = .2959). Under the control condition, pre was 4.23 ± 0.76 Nm/kg and post was 4.04 ± 0.54 Nm/kg (*P* = .1868). Under the PAP condition, pre was 4.16 ± 0.66 Nm/kg and post was 4.35 ± 0.68 Nm/kg (*P* = .0534), showing the highest value in post under the PAP condition.

**Figure 5:**
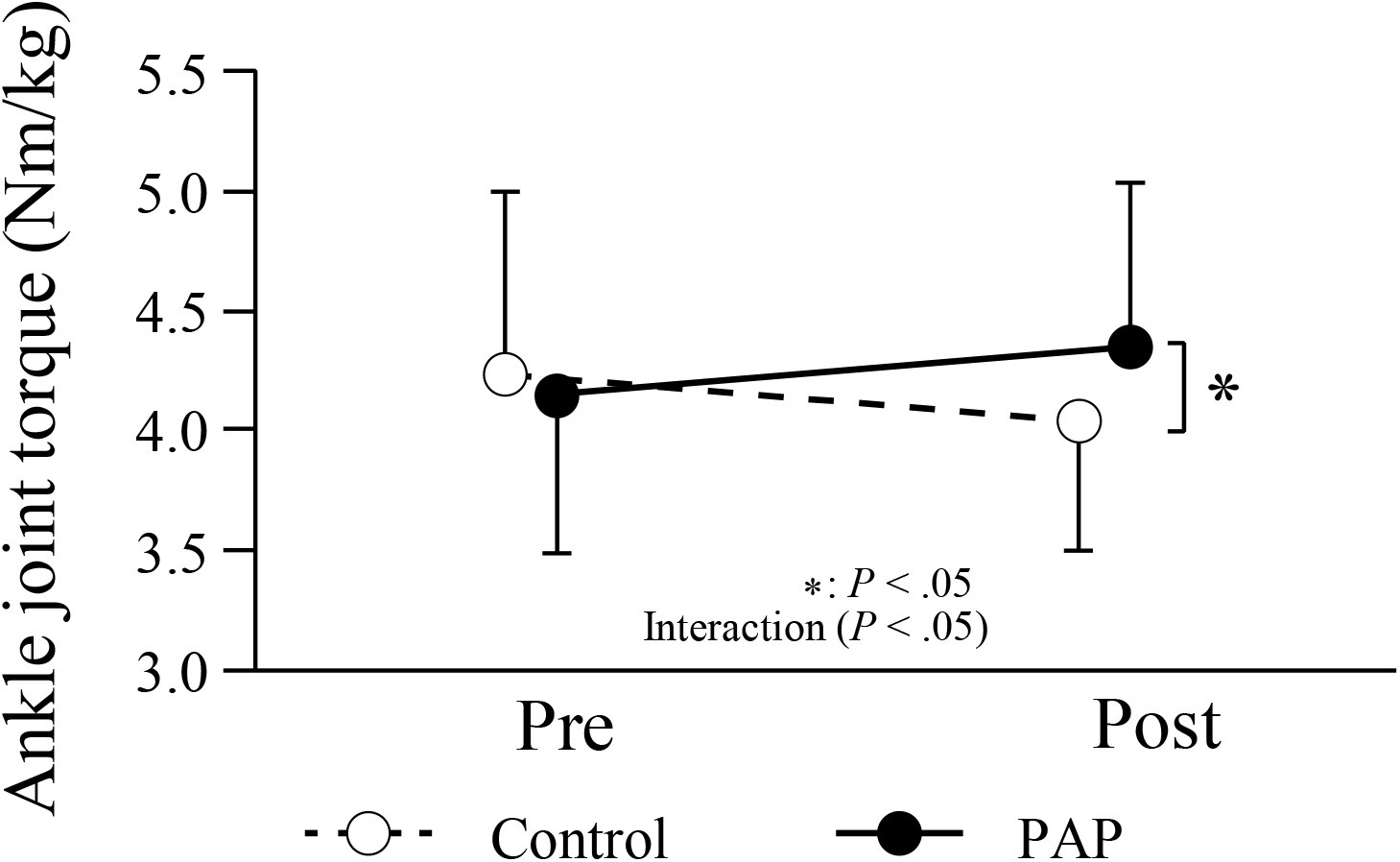
Ankle joint torque in pre and post in PAP and control condition. Circles and error bars indicate means and SDs respectively. Black circle with black line illustrates PAP condition. White circle with dashed line shows control condition.

### Percentage root mean square

Figure 6 shows the %RMS values for each muscle during the descending and ascending phases. No significant interaction was observed in any trials under either condition in all target muscles (TA: *P* = .813 and .51, SOL: *P* = .295 and .0264, MG: *P* = .0604 and .311, LG: *P* = .0942 and .0566, RF: *P* = .674 and .257, BF *P* = .127 and .862, respectively, for the descending and ascending phase) except for SOL in the ascending phase; however, there was no significant difference between any trial (pre vs. post under PAP: *P* = .0754, pre vs. post under control: *P* = .0637, pre vs. pre between conditions: *P* = .1106, post vs. post between conditions: *P* = .0640). The %RMS of the descending phase exceeds 200% for the SOL, MG, LG, and RF. This is because the extensor muscles of the ankle and knee joints antagonize dorsiflexion and flexion movements when each joint in the descending phase increases its angle in the direction of dorsiflexion, causing eccentric contraction. After exceeding the descending phase, the TA and BF, extensor antagonists, generated <80%. The SOL, MG, LG, and RF values were approximately 200% in the ascending phase.

**Figure 6:**
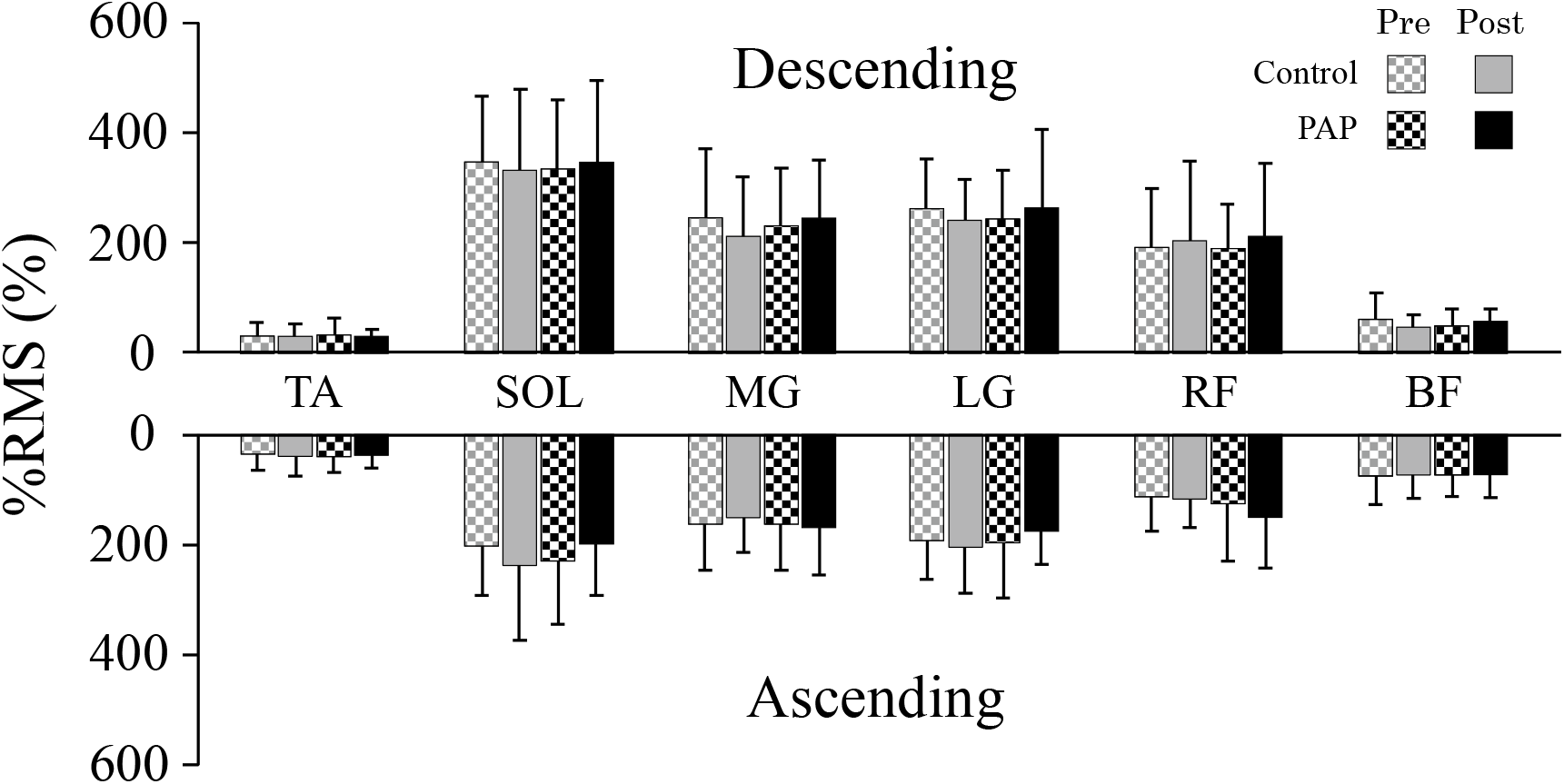
Values of %RMS in pre and post under PAP and control condition in the descending and ascending phases in each muscle. On SOL, interaction was observed in ascending phase but no significant difference between all trials. The values in upper and lower of this graph are divided respectively the descending and ascending phases. Pre in both conditions is illustrated as bars with block pattern, and bars of post in both condition is with no pattern. PAP is colored with black, and control is with gray. Target muscles are written by abbreviations: TA (tibialis anterior), SOL (soleus), MG (medial gastrocnemius), LG (lateral gastrocnemius), RF (rectus femoris), and BF (biceps femoris).

## DISCUSSION

This study had two important points: increased jump performance after CC and the invariance of EMG results between conditions. The jump height of the drop jump increased by 11% (0.04 m); it was realized by the increment in the velocity of the center of gravity at takeoff (0.14 m/s, *P* = .002) and the ground reaction force (11 N s, *P* = .004) in the same ground contact time at post in the PAP condition compared to pre. This increase in performance was caused by the potentiation of ankle torque induced by CC on the triceps surae muscle. However, no difference was observed in muscle activity based on EMG, even in trials with ankle torque potentiation. Consequently, increased jump performance can be assumed because of changes in excitation-contraction coupling. This phenomenon is thought to be caused by a mechanism involving the PAP. This is considered to be due to an increase in the sensitivity of actomyosin to calcium ions due to phosphorylation of the myosin light chain and not the enhancement of muscle strength due to the excitement of the nervous system.^28^ Studies using the same CC^12,15^ showed that non-voluntary muscle contractions evoked by a fixed magnitude of stimulation were potentiated due to PAP. Additionally, a previous study where PAP was observed on drop jump^29^ showed that drop jump height increased with no increment in EMG activity after plyometric exercise inducing PAP on ankle plantar flexion. Herein, PAP on the triceps surae muscle enhanced ankle torque, improving jump height with no significant difference in EMG; similarly, potentiated muscle contraction increased the work of the typical SSC on the ankle joint. Furthermore, if CC can induce PAP in a target muscle, it is not necessarily similar to the target movement and style of muscle contraction, as reported in a previous study. Based on the above findings, CC aids in forming cross-bridges, increases force exertion at the triceps surae muscle, and improves SSC function.

For SSC, the main benefit of strengthening cross-bridge induced by intense muscle activation is preventing cross-bridge dissociation and filament sliding,^14^ which absorb energy provided by eccentric contraction.^30^ Additionally, the cross-bridges are maintained with a small length change for a short time.^6^

In the drop jump situation, when the recruitment of the triceps surae muscle is high, ankle joints can endure the high eccentric load.^19^ Cross-bridge dissociation becomes less likely to occur even during the descending phase of the drop jump as the number of bonds increases and the bonds become stronger. In addition, the behavior of a viscoelastic body is considered to be due to the number of bonds in the cross-bridges;^6^ therefore, this improvement strengthens the SSC. Therefore, the torque of the ankle joints, whose magnitude at the time of switching from the descending phase to the ascending phase reflects the amount of stored elastic energy,^19^ increased. PAP showed an enhancing effect on the ankle joint and the SSC on the triceps surae muscle, not only for simple force exertion.

Therefore, herein, it was suggested that the increase in ankle joint torque with no increment in % RMS occurred due to the CC, which may induce PAP in the triceps of the lower leg. PAP induced only on the triceps surae muscle increased the work of the ankle joint by reducing cross-bridge dissociation in the descending phase of the drop jump, which improved jump height. To increase the function of the SSC by PAP, it is not necessary to have a similar movement or style of muscle contraction if PAP can be induced in the target muscle. The results confirmed the importance of the ankle joint in jump performance.

## PRACTICAL APPLICATIONS

This result suggests that the CC may be used to improve SSC function for muscles involved primarily in any movement where SSC occurs. Furthermore, this CC is easier to apply to further researches for PAP and pre-exercise for impulsive movements than to high-load CC because it causes less fatigue, requires no rest time, and has fewer individual differences. Furthermore, this study demonstrated that increasing the ankle torque of plantarflexion was strengthened by the potentiation of triceps surae muscle contraction while jump height of drop jump increased. Therefore, it indicates the importance of the training on the muscle for plantarflexion for jump performance.

## CONCLUSIONS

This study investigated the possibility of using PAP to improve SSC function in ankle joints. To induce PAP on the triceps surae muscle, maximal voluntary isometric plantar flexion was performed for 6 s as CC and 10 s before the drop jump. Performing CC, which only activates the triceps surae muscle, increased the ankle torque of plantar flexion and increased the jump height and its relatives: the velocity of the center of gravity at takeoff and ground reaction force. CC is unnecessary to have a similar movement or style of muscle contraction to target movement for increasing jump performance, such as a drop jump, which SSC mainly aids, if PAP can be induced in the target muscle by CC as demonstrated in this study. Furthermore, the results confirmed the impact of ankle joints and the triceps surae muscle on jump performance.

## Data Availability

All data produced in the present work are contained in the manuscript

## ACKNOWLEDGMENTS

The authors thank the athletes and coaches of the Track and Field clubs for their time. The authors declare no conflicts of interest. The results of the current study do not constitute endorsement of the product by the authors or the journal.

